# From Policy Change to Program Impact: HPV Vaccination Coverage Following the Global Shift to a Single-Dose Schedule

**DOI:** 10.64898/2026.11.19.26368547

**Authors:** Marina Terada, Ping Teresa Yeh, Helen Natalia, Lucy Hribar Baker, Mauricio Torres-Martinez, Damilola Bolanle, Humyra Shah, Uswa Jiwani, Leanne Zhang, Soim Park

## Abstract

**Background:** Since the World Health Organization (WHO) endorsed single-dose HPV vaccination in 2022, many countries have adopted this schedule. However, evidence regarding the schedule changes’ impact on HPV vaccination coverage remains limited. Using a sequential explanatory mixed-methods design, we assessed coverage changes following the schedule switch and explored stakeholders’ perspectives on factors influencing these trends.

**Methods:** Using WHO-UNICEF Estimates of National Immunization Coverage, we assessed first-dose HPV vaccination (HPV1) coverage among females in 49 countries that switched to a single-dose schedule during 2022-2024. We used Wilcoxon signed-rank tests to compare mean HPV1 coverage before and after the switch, stratified by country and program characteristics. We also examined country-level changes in HPV1 and complete HPV vaccination (HPVc) coverage across all 49 countries. We then qualitatively interviewed 33 national and sub-national HPV vaccination stakeholders from 15 countries and analyzed the data using thematic analysis.

**Results:** Following the switch, significant increases in mean HPV1 coverage were observed among non-high-income countries (55.0% pre-switch vs. 63.3% post-switch, p=0.019), countries in the WHO African Region (58.9% vs. 71.6%, p=0.038), and programs employing school-based delivery (57.0% vs. 66.0%, p=0.010). Significant increases were also observed in programs targeting females only (56.3% vs. 65.9%, p=0.040), those targeting single-age cohorts (61.3% vs. 72.0%, p=0.013), and Gavi-supported countries (54.9% vs. 73.2%, p=0.010). Stakeholders attributed coverage trends to multiple factors beyond the schedule change, including recovery from COVID-19-related disruptions, concurrent programmatic changes, and broader contextual factors.

**Conclusion:** Although coverage trends varied across countries, improvement in HPV1 coverage in resource-limited settings suggested that the switch to a single-dose schedule may have facilitated recovery from COVID-19-related disruptions. Continued monitoring of vaccination coverage, implementation outcomes, and program sustainability is essential to understanding the long-term population-level implication of the single-dose recommendation.

## Introduction

Cervical cancer is the fourth most common cancer in incidence and mortality among women, with an estimated 660,000 new cases and 350,000 deaths in 2022 globally [1]. In 2020, the highest numbers of incidence and deaths were reported in Asia and Africa regions [2]. Vaccination against human papillomavirus (HPV) is critical in preventing high-risk HPV16 and HPV18 infections, to which 77% of the global burden of cervical cancer is attributed [3]. In 2020, the World Health Organization (WHO) proposed a set of global cervical cancer elimination goals by 2030, including 90% of girls fully vaccinated with HPV vaccine by the age of 15 [4]. Since then, global coverage among females receiving at least one dose gradually increased from 17% in 2019 to an estimated 33% in 2025 [5]. However, significant disparities in uptake persist across countries, particularly when stratifying by World Bank income classification [6].

In 2022, the WHO’s Strategic Advisory Group of Experts (SAGE) on immunization updated its recommendations for HPV vaccination, shifting from previous multiple-dose regimens to a single- or two-dose schedule for girls aged 9–20 years old based on current evidence of efficacy trials [7,8]. A single-dose of the HPV vaccine demonstrated comparable efficacy and effectiveness against HPV infection to a two-dose regimen, showing noninferiority in preventing high-risk strains of HPV among girls [9–12]. This protection was sustained for at least 10 years of follow-up [13]. Following the WHO SAGE recommendations and review of this scientific evidence, an increasing number of countries either switched to a single-dose schedule or introduced HPV vaccination using a single-dose regimen [14], with 19 Gavi-supported HPV vaccine introductions in 2023-2024 adopting a single-dose schedule [15].

Introducing a single-dose schedule was expected to offer programmatic advantages and benefits, including greater implementation efficiency, reduced financial and economic costs [16], and improved coverage [17]. A qualitative study with national immunization stakeholders in non-high-income countries (HICs) highlighted several motivators for adopting a single-dose HPV vaccination schedule in resource-limited settings [17–19], including financial and logistical benefits, improved program performance, and a greater public health impact in reducing the burden of cervical cancer, particularly among hard-to-reach populations.

However, the impact of switching to a single-dose regimen on HPV vaccination coverage and the contextual factors underlying those changes remain under investigated. A total of 96 countries have adopted this regimen as of August 2026 [14]. Despite its growing adoption, only a small number of studies have to date assessed the actual or anticipated advantages and disadvantages of the switch [17,20]. Additionally, a previous global analysis of HPV vaccine coverage reported a limitation of the lack of country-specific background information needed to further explore underlying reasons for coverage trends [21]. Thus, we conducted a mixed-methods study with a sequential explanatory design to: 1) quantitatively assess changes in HPV vaccination coverage before and after countries switched from a two-dose to a one-dose HPV vaccination schedule, and 2) qualitatively explore the country-specific contexts that may have influenced the post-switch coverage trends.

## Methods

### Study Design

Employing a sequential explanatory mixed-methods design [22,23], we first quantitatively investigated the changes in HPV vaccination coverage among 49 countries that had switched to a single-dose schedule by the end of 2024, using WHO-UNICEF Estimates of National Immunization Coverage (WUENIC) data published in July 2025 [24]. Based on observed coverage trends following the schedule switch, we classified countries into three groups: increased, decreased, or stable which we operationally defined as a change within 5 percent points. We selected 15 countries for inclusion in the qualitative phase, prioritizing variation in geography, country income level, and post-switch HPV1 coverage trends. We then conducted key informant interviews with national and sub-national HPV vaccination stakeholders in these countries to explore their perspectives on the schedule switch and identify internal and external factors that may have contributed to the change in vaccination coverage. The countries included in the study are shown in **Figure 1**.

**Figure 1.**
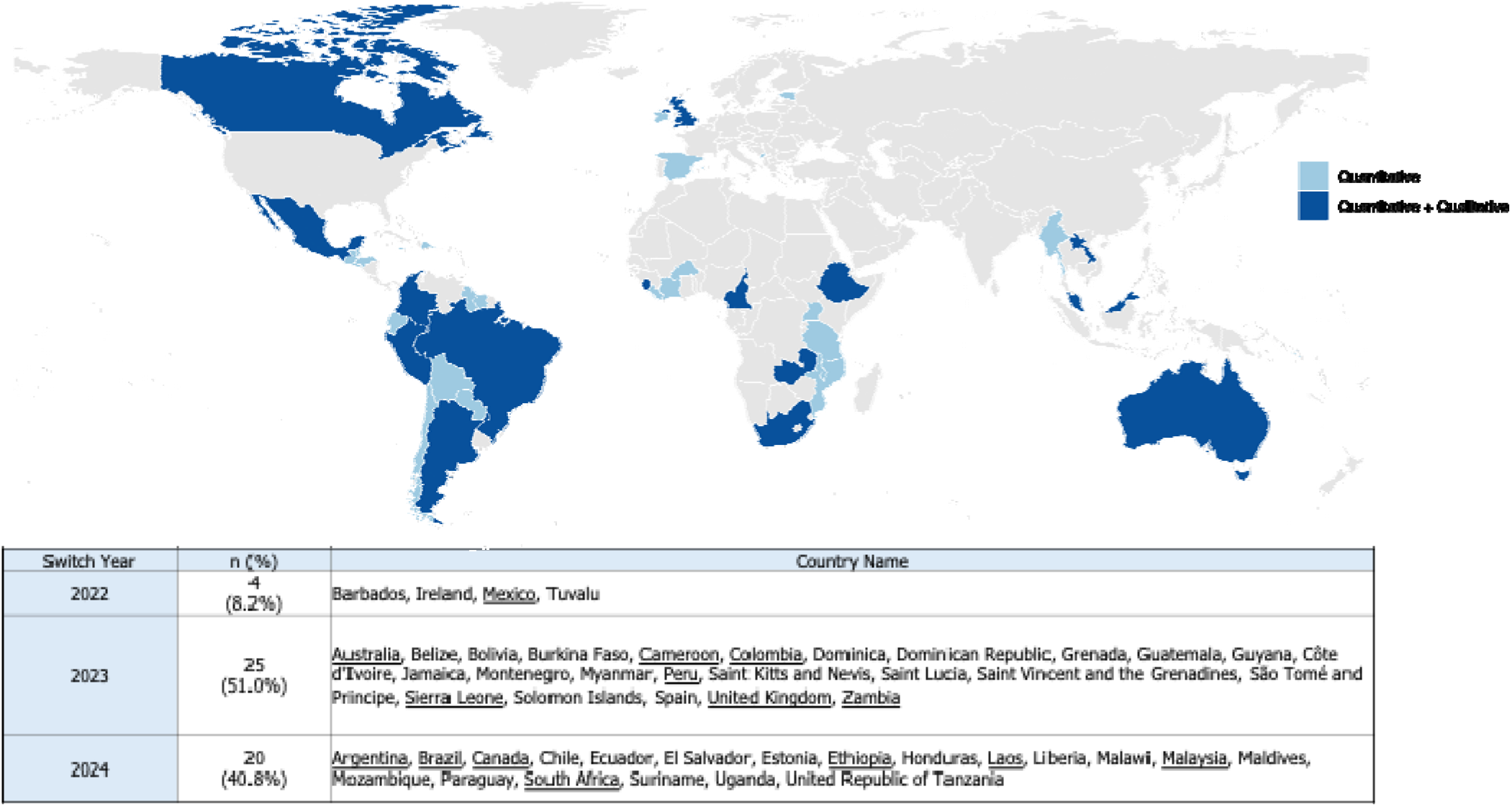
World map of 49 countries showing the year of transition to a single-dose HPV vaccination schedule and the analytical methods used. Countries with underlined labels were included in both quantitative and qualitative analyses (n = 15). Barbados, Dominica, Grenada, Saint Kitts and Nevis, Saint Lucia, Saint Vincent and the Grenadines, Sao Tome and Principe, and the Maldives are not visible due to their small land area at the scale used in this map.

### Data Sources and Measures

We obtained vaccination coverage data from the publicly available WUENIC 2025 [25], which provides annual estimates of HPV vaccination coverage among the girls targeted by national vaccination programs in each calendar year through 2024 [24]. The HPV vaccination coverage estimates are based on annual administrative and survey data submitted by each country to WHO through the Joint Reporting Form on Immunization. Specifically, we extracted two program coverage estimates for females in each country: national program coverage for the first dose of HPV vaccination (HPV1) and for completion of the full HPV vaccination (HPVc) series.

We also obtained information on each country’s HPV vaccination program characteristics from the WHO HPV Dashboard and the VIEW-hub by the International Vaccine Access Center [14,26]. These characteristics included the primary delivery strategy (e.g., school-based, facility-based, or mixed delivery), target age cohort, target sex, and Gavi support status. Although the WHO HPV dashboard provides information on the current dose regimen for each country, it does not indicate the year when each country switched its program. Therefore, we first determined the year of switch from multiple doses to a single dose based on the WHO HPV Dashboard [14], identifying the year when HPV1 and HPVc coverage became identical. We further confirmed the switched year for each country through consultation with the WHO and stakeholders from relevant countries. Only countries that switched to a single-dose HPV vaccination program from 2022 to 2024 were included in our analysis.

### Quantitative Analysis

First, we identified countries that have switched to single-dose programs in 2022, 2023, or 2024, and conducted a descriptive analysis of each country’s HPV vaccination program characteristics. To assess the coverage trends, we calculated population-adjusted coverage for a single-age cohort, grouping countries by the HICs and non-HICs based on the World Bank’s income classification at the time of analysis [27]. To account for variation in target age at vaccination across countries (9 to 14 years), we standardized estimates using the population of 15-year-old girls as the single-age cohort, as recommended by the WHO [24]. For each country and year, the estimated number of vaccinated girls was calculated by multiplying the HPV1 or HPVc coverage by the corresponding population of 15-year-old girls. Age- and sex-specific population estimates were obtained from Estimated population data from the United Nations (UN) Population Division [28]. Because 2024 population estimates were unavailable at the time of analysis, we used the medium-variant population projection, which assumed mean fertility and mortality and median net migration for projections [29]. The formula when calculating the population-adjusted coverage is:

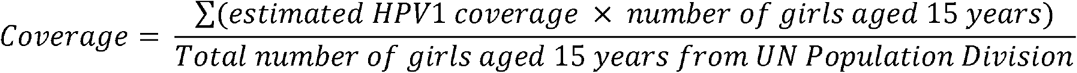

To assess group-specific changes in HPV1 coverage before and after the switch to a single-dose schedule, we performed Wilcoxon signed-rank test using paired country-level mean values across six country and program characteristics, namely World Bank income classification, WHO region, primary delivery strategy, target sex, target age cohort, and Gavi support status. Exact p-values were calculated when subgroup sample sizes were small. We visualized pre- and post-switch coverage distributions using box plot graphs. Years with missing coverage data were excluded from the calculation of country-level means, and coverage estimates truncated at 99% were excluded following the convention used in a previous study [6]. For country-level changes in coverage, mean HPV1 and HPVc coverage was calculated for the pre-switch period (national introduction through the year before the switch) and the post-switch period (the year of the switch through 2024). The results were presented in a bubble chart.

As a sensitivity analysis, we repeated country-level changes in coverage analyses treating 2020-2021 separately to account for potential disruptions in HPV vaccination coverage associated with the COVID-19 pandemic, which disproportionately affected coverage in non-HICs compared to HICs [21]. We used a two-tailed significance level of α=0.05 and conducted all analyses using R version 4.5.2 (R Core Team, 2025, Vienna, Austria).

### Stakeholder Interviews

Based on the quantitative results, countries were categorized as having increased, stable, or decreased coverage following the switch and further grouped by World Bank classification and Gavi support status (**S1 Table**). From each category, we purposively selected two or three countries. Eligible key informants were national- or regional-level stakeholders with professional involvement in HPV vaccine implementation or policy, including representatives from Ministries of Health, Gavi-supported technical assistance partners, multilateral organizations, civil society organizations and academic institutions. Initial participants were identified through the HPV vaccine Acceleration Program Partners Initiative (HAPPI) Consortium networks, with additional participants recruited via snowball sampling. Of the 49 countries that switched to a single-dose schedule during 2022-2024, we contacted 84 stakeholders from 19 countries. Between October 2025 and March 2026, we conducted 33 interviews with stakeholders from 15 countries, including three HICs (Australia, Canada, and the United Kingdom), seven upper-middle-income countries (UMIC; Argentina, Brazil, Colombia, Malaysia, Mexico, Peru and South Africa), three lower-middle-income countries (LMICs; Cameroon, Laos and Zambia), and two low-income countries (LICs; Ethiopia and Sierra Leone).

We developed a semi-structured key informant interview guide to explore experiences with single-dose schedule adoption and implementation, including questions on HPV vaccination coverage trends (see **S2 Table** for relevant questions). During the interviews, we presented country-specific coverage trends from the quantitative analysis and invited participants to interpret the observed patterns and identify potential contributing factors. The qualitative findings presented in this manuscript were largely drawn from these discussions. Interviews were conducted virtually by nine researchers with graduate-level training in qualitative research methods. To minimize language barriers, key informant interviews were conducted in English, Spanish, Portuguese, or French. Participants were interviewed individually via an audio-conferencing platform for approximately 30 to 60 minutes.

### Qualitative Analysis

All interviews were audio-recorded after oral consent and transcribed verbatim using the transcription feature in Microsoft Word for the Web. Non-English transcripts were translated into English using DeepL and ChatGPT. All transcripts were quality-checked for transcription and translation accuracy by the original interviewer or another research team member. We conducted a team-based thematic analysis to identify patterns across interviews and generate analytic categories related to contextual factors contributing to coverage trends. We developed an analytic framework to evaluate stakeholder perspectives on post-switch coverage trends, considering the timing of the switch in each country. Each transcript was individually coded by one of seven researchers, with one team member providing ongoing oversight of the analytic process. We used Atlas.ti Web version 9.24.3 (ATLAS.ti Scientific Software Development GmbH, Berlin, Germany) to facilitate data management, coding, and interpretation.

### Mixed-Methods Integration

Quantitative and qualitative components were integrated at multiple stages of the study. Findings from the quantitative analysis informed purposive selection of countries for the qualitative component, with the aim of including countries that reflected diverse coverage patterns and program contexts. During interviews, country-specific quantitative findings were presented to participants near the end of the interview, and participants were asked to interpret the observed coverage trends and describe contextual factors that may have contributed to them. The qualitative findings were then used to contextualize and help explain the quantitative results, including similarities and differences in coverage changes across countries.

### Ethical Approval

The Johns Hopkins Bloomberg School of Public Health Institutional Review Board determined that the study was exempt from human subjects research oversight. Oral informed consent was obtained from all key informants.

## Results

### Country and Program Characteristics

Among the 49 countries that switched to a single-dose schedule between 2022-2024 (**Table 1**), most were classified as non-HICs (79.6%, n=39). School-based delivery strategies were the most common approach (75.5%, n=37). Nearly half of the countries (44.9%, n=22) targeted females only. Single age cohorts accounted for the largest proportions (57.1%, n=28), while other countries targeted multiple age cohorts (38.8%, n=19). Gavi-supported countries accounted for 30.6% (n = 15).

**Table 1.** Characteristics of HPV vaccination programs in 49 countries that switched to a single-dose program during 2022-2024.

| Characteristic | Category | n (%) |
|---|---|---|
| Switch Year | 2022 | 4 (8.2) |
|  | 2023 | 25 (51.0) |
|  | 2024 | 20 (40.8) |
| World Bank Income Level | HICs | 10 (20.4) |
|  | Non-HICs^ | 39 (79.6) |
| WHO region | African Region | 13 (26.5) |
|  | Region of the Americas | 24 (49.0) |
|  | European Region | 5 (10.2) |
|  | South-East Asia Region | 2 (4.1) |
|  | Western Pacific Region | 5 (10.2) |
| Primary HPV Vaccine Delivery Strategy | School-based | 37 (75.5) |
|  | Facility-based | 3 (6.1) |
|  | Mixed | 5 (10.2) |
|  | Varies by region/province | 1 (2.0) |
| Target Sex | Male and Female | 27 (55.1) |
|  | Females only | 22 (44.9) |
| Target Age Cohort | Single Age Cohort | 28 (57.1) |
|  | Multiple Age Cohort | 19 (38.8) |
| Gavi Status | Gavi Country | 15 (30.6) |
|  | Non-Gavi Country | 34 (69.4) |

Note: Some countries lacked information on the target age cohort or primary delivery strategy; therefore, totals may not sum to 49.
^ Non-high-income countries (non-HICs) include low-income countries (LICs), lower-middle-income countries (LMICs), and upper-middle-income countries (UMICs).

### Population-Adjusted Trends in HPV Vaccination Coverage

Population-adjusted HPV1 coverage among girls aged 15 in non-HICs showed a decline during the COVID-19 pandemic, dropping from 59.6% in 2019 to 45.7% in 2021 (**Figure 2**). In contrast, coverage in HICs showed a stable trend, remaining stable at above 80% during the same period. In 2024, 73.6% of girls aged 15 years in non-HICs that switched to a single-dose schedule (n=38) received one dose, compared to 81.5% of girls in HICs (n=10) who also received one dose. Population-adjusted HPVc coverage, defined as the proportion of girls in the target population who received all doses required under the national schedule, among girls aged 15 in non-HICs showed a more drastic decline during the COVID-19 pandemic, dropping from 59.7% in 2019 to 31.9% in 2021 (**S3 Figure**).

**Figure 2.**
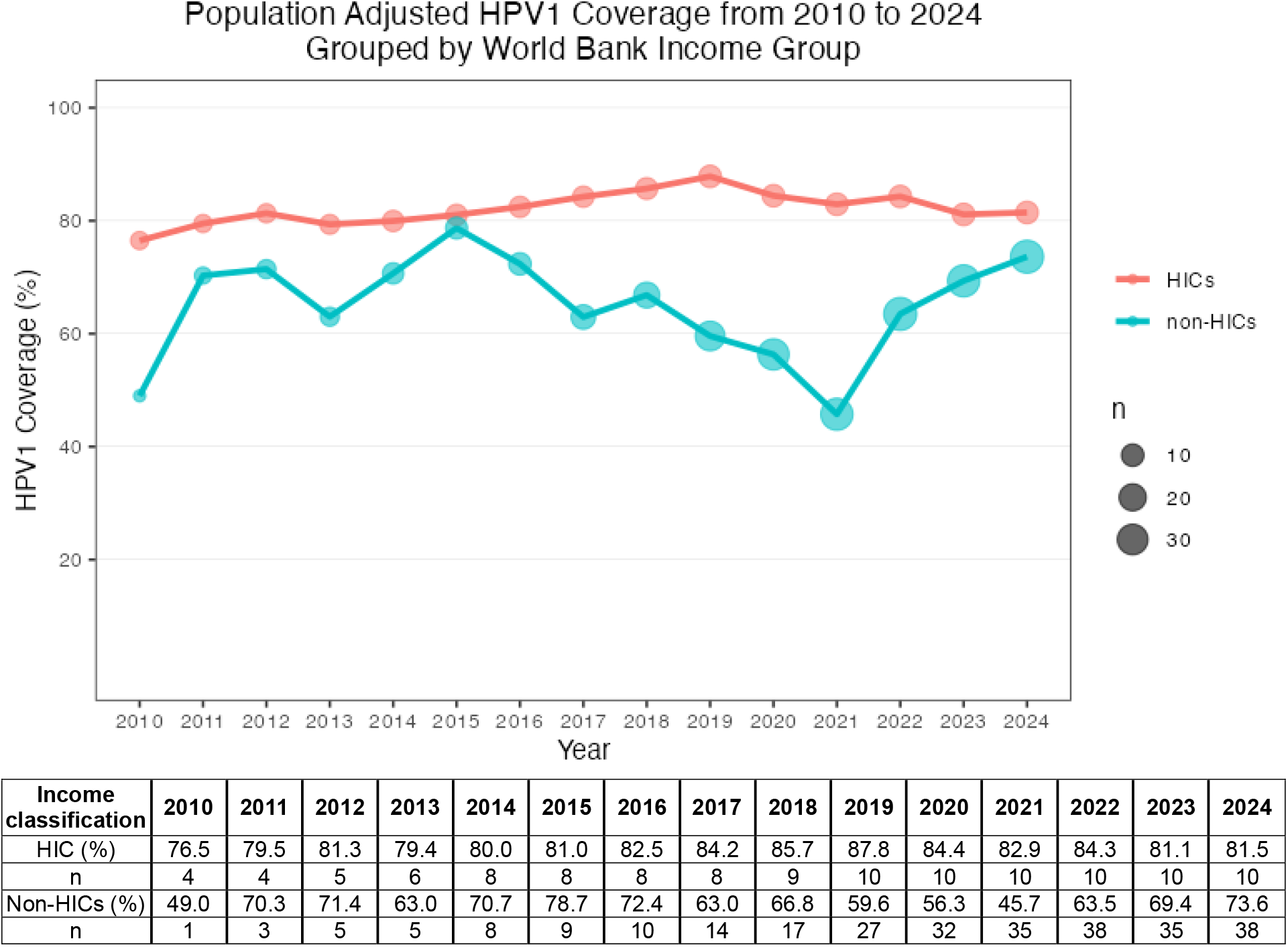
Population-adjusted HPV1 vaccination coverage from 2010 to 2024, grouped by World Bank income category. Note: Implausible coverage estimates (i.e., 99%) were excluded from the analysis. Country income classifications were based on the 2025 World Bank income categories. Non-high-income countries (non-HICs) include low-income countries (LICs), lower-middle-income countries (LMICs), and upper-middle-income countries (UMICs).

### Pre- and Post-Switch Coverage by Country and Program Characteristics

When grouped by World Bank income classification, non-HICs significantly increased their mean HPV1 coverage from 55.0% before the switch to 63.3% after the switch (p=0.019), whereas HICs showed a non-significant increase in mean HPV1 coverage from 70.3% to 73.0% (Table 2, **S4 Figure**). Countries in the African region also showed an increase, rising from 58.9% before the switch to 71.6% after the switch (p=0.038). Countries implementing school-based delivery increased their coverage from 57.0% before the switch to 66.0% after the switch (p=0.010). Additionally, countries targeting only females increased coverage from 56.3% to 65.9% (p=0.040), those targeting a single age cohort increased from 61.3% to 72.0% (p=0.013), and Gavi-supported countries went from 54.9% to 73.2% (p=0.010).

**Table 2.** Mean HPV1 coverage based on World Bank income classification, WHO region, primary delivery strategy, target sex, target age cohort and Gavi status.

| **Characteristic** | **Category** | n^ | HPV1 coverage (%) |  |  |
|---|---|---|---|---|---|
|  |  |  | Pre-switch | Post-switch | p-value |
| World Bank Income Classification (binary) | HICs | 10 | 70.3 | 73.0 | 0.959 |
|  | Non-HICs^ | 39 | 55.0 | 63.3 | **0.019** |
| WHO Region | Region of the Americas | 24 | 55.6 | 58.4 | 0.584 |
|  | African Region | 13 | 58.9 | 71.6 | **0.038** |
|  | Other regions<sup>†</sup> | 12 | 62.5 | 72.3 | 0.255 |
| Primary Delivery Strategy | School-based | 37 | 57.0 | 66.0 | **0.010** |
|  | Facility-based/Mixed | 8 | 54.4 | 63.4 | 0.363 |
| Target Sex | Both sexes | 27 | 59.7 | 64.8 | 0.248 |
|  | Females only | 22 | 56.3 | 65.9 | **0.040** |
| Target Age Cohort | Single Age Cohort | 28 | 61.3 | 72.0 | **0.013** |
|  | Multiple Age Cohort | 19 | 51.5 | 54.2 | 0.695 |
| Gavi Support Status | Gavi Country | 15 | 54.9 | 73.2 | **0.010** |
|  | Non-Gavi Country | 34 | 59.6 | 61.8 | 0.538 |

Note: Coverage analyses included the COVID-19 period and excluded implausible coverage estimates (i.e., 99%).
Sums may not equal 49 due to missing data. For each country, mean HPV1 and HPVc coverage were calculated for the pre-switch period (2010 to the year before the switch) and the post-switch period (from the switch year through 2024).
^ Non-high-income countries (HICs) include low-income countries (LICs), lower-middle-income countries (LMICs), and upper-middle-income countries (UMICs).
<sup>†</sup> Combined three regions (European, South-East Asian, and Western Pacific Regions) due to small sample size.
Abbreviations: HIC: High-income country; WHO: World Health Organization.

### Country-Level HPV Vaccination Coverage Changes Before and After the Switch

Regarding country-level coverage trend (**Figure 3**), the mean HPV1 coverage among HICs (n=10), including Australia, Barbados, Saint Kitts and Nevis, and the United Kingdom, indicated a decrease following the switch. In contrast, Ireland’s coverage remained relatively stable, while Canada, Chile, Guyana, Estonia and Spain experienced an increase. Among UMICs, Argentina, Brazil, Dominica, Grenada, Jamaica, Montenegro, Paraguay, Saint Vincent and the Grenadines, Suriname and Tuvalu showed a decrease in coverage. Additionally, among LICs and LMICs receiving Gavi support, Sierra Leone, and Zambia also demonstrated a decrease in HPV1 coverage.

**Figure 3.**
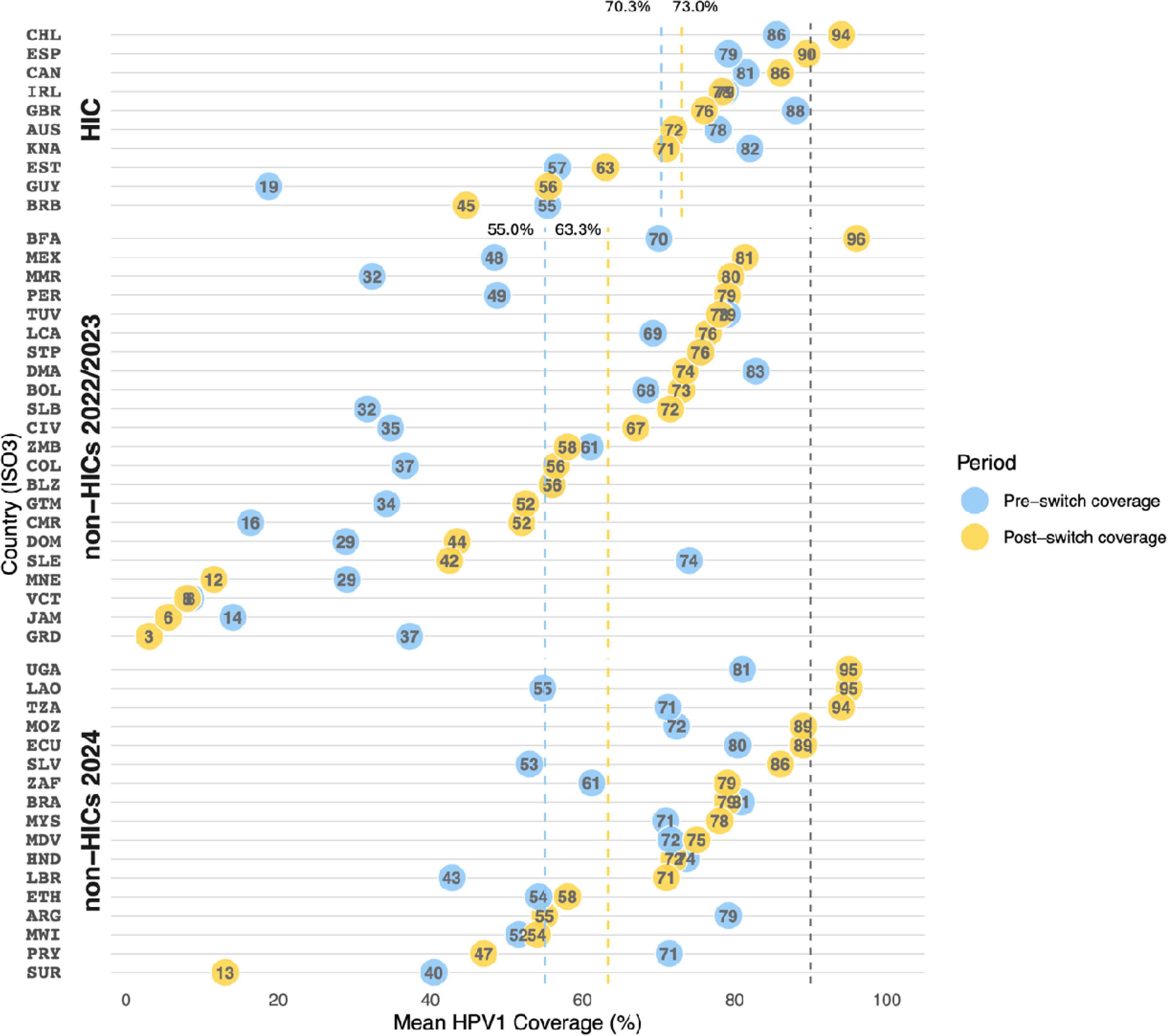
Bubble chart showing mean HPV1 coverage before and after the switch to a single-dose schedule among high-income countries (HICs) and non-HICs. Note: Implausible coverage estimates (i.e., 99%) were excluded from the analysis. Countries were grouped into high-income countries (HICs) and non-HICs, as well as by the year of the switch (2022–2023 vs. 2024). Country names in the figure are presented using ISO Alpha-3 country codes. The dotted black line represents the WHO target of 90% HPV vaccination coverage, and the colored dotted lines represent the mean coverage levels, with yellow indicating post-switch coverage and blue indicating pre-switch coverage for HICs and non-HICs. For Belize (BLZ) and São Tomé and Príncipe (STP), the pre- and post-switch mean coverage values were identical; therefore, the pre- and post-switch bubbles completely overlap and appear as a single bubble.

Country-level trends in mean HPVc coverage are presented in **Figure 4**. Most countries recorded higher HPV vaccination completion coverage following the single-dose switch in both HICs and non-HICs. From a programmatic perspective, the substantial increases in completion coverage indicate that national programs achieved their vaccination objective for a larger proportion of eligible girls following the single-dose switch.

**Figure 4.**
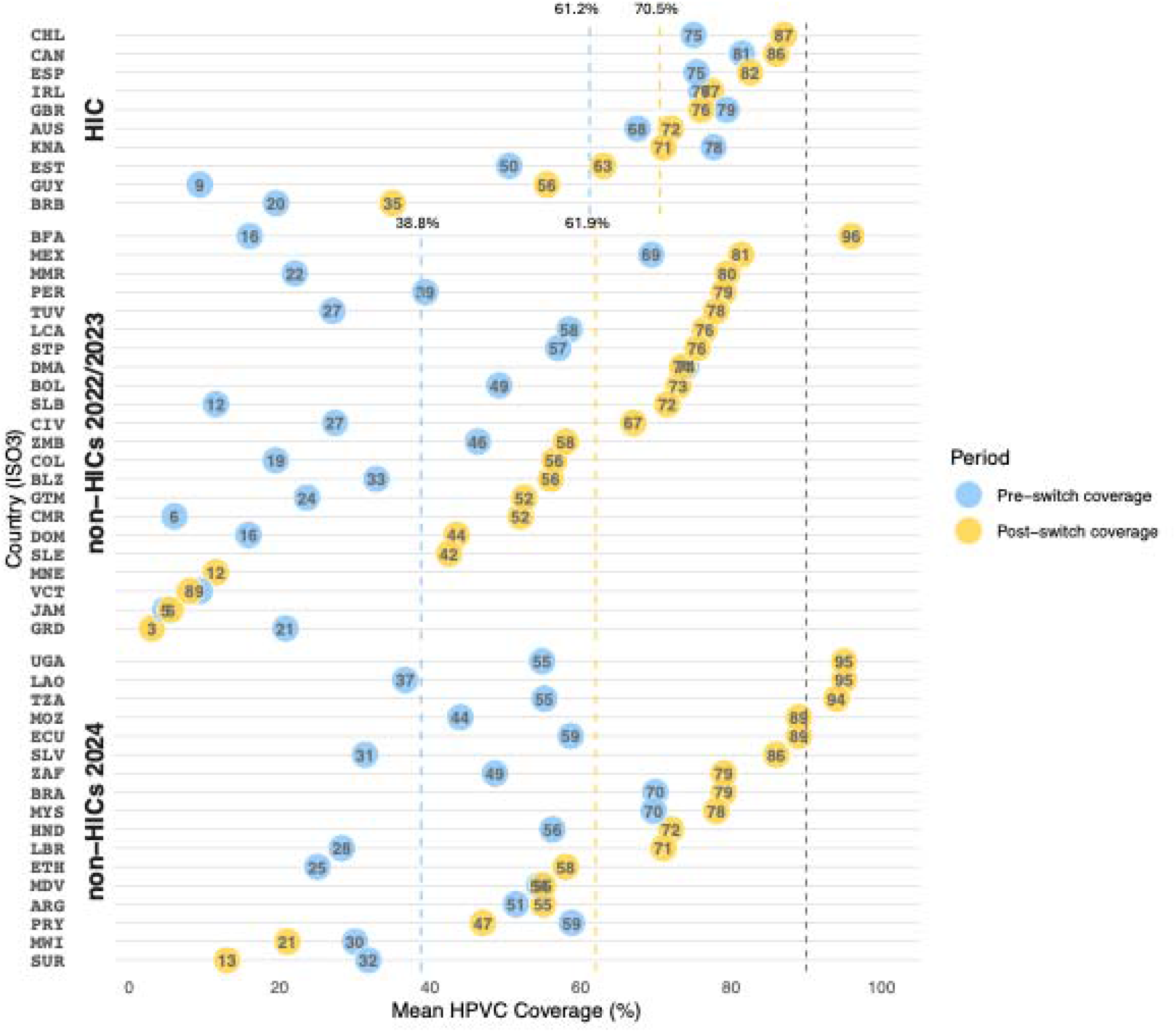
Bubble chart showing mean HPVc coverage before and after the switch to a single-dose schedule among high-income countries (HICs) and non-HICs. Note: Implausible coverage estimates (i.e., 99%) were excluded from the analysis. Countries were grouped into high-income countries (HICs) and non-HICs, as well as by the year of the switch (2022–2023 vs. 2024). Country names in the figure are presented using ISO Alpha-3 country codes. The dotted black line represents the WHO target of 90% HPV vaccination coverage, and the colored dotted lines represent the mean coverage levels, with yellow indicating post-switch coverage and blue indicating pre-switch coverage for HICs and non-HICs. Mean post-switch coverage differs between HPV1 in Figure 3 and HPVc in Figure 4 because some countries reported different HPV1 and HPVc coverage estimates for these indicators in the switch year. For Montenegro (MNE) and Sierra Leone (SLE), pre-switch HPVc coverage data were unavailable; therefore, only the post-switch coverage is shown.

Results of sensitivity analyses examining coverage estimates separately during the COVID-19 pandemic are shown in **S5 and S6 Figures**. In HICs, coverage levels before and during the COVID-19 pandemic were largely similar. In contrast, among the non-HICs that introduced HPV vaccination programs before the pandemic, some Gavi-supported countries (e.g., Côte d’Ivoire, Tanzania, and Liberia) experienced declines in coverage during the pandemic, which may have contributed to lower pre-switch coverage. Overall, non-HICs tended to exhibit lower HPV completion coverage than HICs, with data points concentrated at lower coverage levels.

### Stakeholder Perspectives on Factors Shaping Coverage

Among the 33 qualitative study participants, 18 (54.5%) were affiliated with Ministries of Health or Expanded Programmes on Immunization, followed by nonprofit organizations such as technical assistant partners, non-governmental organizations, and civil society organizations (n=9, 27.3%), and academic institutions (n=4, 12.1%). Two participants (6.1%) were affiliated with both academic and governmental institutions. On average, we interviewed 2.2 stakeholders per country with the number ranging from 1 to 4.

Participants expressed mixed reactions to the country-specific coverage trends presented during the interviews. Most considered the findings broadly consistent with their expectations, but some noted discrepancies between the WUENIC estimates and their national data. Across countries, stakeholders cautioned against attributing changes in HPV vaccination coverage solely to the switch to a single-dose schedule. They emphasized that the switch occurred during recovery from COVID-19-related disruptions, concurrent changes to HPV vaccination programs, political and economic shifts, and other contextual factors.

Countries that experienced a decline in HPV vaccine coverage attributed them primarily to the COVID-19 pandemic rather than to the switch itself. In non-HICs, reported disruptions included vaccine stockouts; vaccine fatigue; increased vaccine hesitancy, misinformation and disinformation; and reduced trust in public health institutions, governments, and healthcare provider recommendations. Although several UMICs and HICs were able to sustain school-based HPV vaccination programs during the pandemic, school closures and persistently low school attendance reduced opportunities to reach adolescents. Stakeholders also noted that, under the previous multi-dose schedule, subsequent school visits provided additional opportunities to vaccinate adolescents who had missed an earlier session, whereas the single-dose schedule could limit these opportunities. Although facility-based vaccination services were available, they were perceived as less effective during the pandemic because many individuals avoided healthcare facilities.

> *We are seeing lower attendance at schools. We haven’t quite bounced back to the attendance rate that we were seeing prior to the pandemic. So, on a given day in a high school, up to 20% of children may be absent. […] One of the things we were very conscious of from a policymaking level was that we had the potential to see a fall in coverage with a single dose because we wouldn’t be visiting a school twice routinely in one year. (Australia)*

In all countries except Australia and UK, the switch to a single-dose schedule also coincided with broader changes to HPV vaccination programs, including adoption of a gender-neutral vaccination, multi-age cohort (MAC) campaigns or catch-up vaccination activities, integration of HPV vaccination into routine immunization services, and expansion of multi-dose HPV vaccination for immunocompromised populations. Stakeholders considered these concurrent programmatic changes, together with the single-dose switch, as contributing to improvements in vaccination coverage. Coinciding with the switch, Malaysia and Zambia temporarily implemented the MAC campaign to address missed cohorts.

> *The switch to a single-dose HPV schedule in 2023 was part of a broader changes to the program and not a stand-alone action. Between 2021 and 2023, Malaysia experienced significant disruptions of HPV vaccine supply, which resulted in several cohorts of girls missing their vaccinations. So, to address this backlog, we had a national catch-up HPV vaccination program [in 2023]. (Malaysia)*

In Cameroon, the expansion of HPV vaccination eligibility to boys improved acceptance of the program among both communities and healthcare workers. Several UMICs and HICs also broadened multi-dose eligibility for immunocompromised populations (e.g., people living with human immunodeficiency virus (HIV) or receiving immunosuppressive therapy), as well as other specific populations, such as victims of sexual abuse and sexual minorities.

Other adaptations that stakeholders identified as impacting coverage included changes in vaccination registration systems, refresher training for health workers, and intensified communication strategies, such as community outreach, mass media campaigns and advocacy activities. In Zambia, a stakeholder attributed the improvement in HPV vaccine coverage to a shift from a campaign-based approach to routine delivery through schools and health facilities:

*“It [coverage change] has nothing to do with the switch. It has to do with routinization. It has to do with the modality of how we deliver the vaccines and how that changed.” (Zambia)*

Stakeholders also identified broader contextual factors that could negatively influence coverage, including vaccine shortages, health worker fatigue, provider reluctance to vaccinate younger girls, and changes in parental consent procedures. Political context affected HPV vaccination coverage as well. For example, in Mexico, despite an overall increase in coverage, stakeholders reported that governmental transitions and resulting changes to vaccine procurement and supply chain systems disrupted program implementation and adversely influenced vaccination efforts. In some countries, strong political commitment, dedicated budget exclusively for HPV vaccination programs, and sufficient resources facilitated the organization of orientation meetings with schools and reach older girls at the middle school level.

> *The country’s political commitment is important because this HPV vaccination has gained credibility and it has been promoted as part of the fight against cancer. It has also been incorporated into the political platform of the actors. An exclusive budget solely for this type of vaccination has helped us a lot. (Peru)*

## Discussion

In this mixed-methods study of 49 countries that switched to a single-dose HPV vaccination schedule by 2024, we found that HPV1 coverage increased when comparing the periods before and after the switch. When stratified by country characteristics, significant HPV1 coverage increase was observed among non-HICs, African region countries, countries targeting female only, countries employing school-based delivery strategies, Gavi-supported countries, while substantial heterogeneity was observed at the country-level. However, these findings do not necessarily indicate that the switch alone was responsible for the observed increases in coverage. Our qualitative findings suggested that coverage trends reflected the combined influence of the schedule switch and other internal and external factors, such as recovery from COVID-19-related disruptions, concurrent programmatic changes to HPV vaccination programs, and political and economic conditions. These findings jointly indicate that switch to a single-dose schedule may have facilitated coverage improvement but was unlikely to have been the sole determinant of coverage trajectories.

Changes to vaccination schedules have traditionally raised concerns about potential declines in vaccine coverage and increased uncertainty among parents and providers. For example, the introduction of the inactivated poliovirus vaccine was anticipated to increase vaccine hesitancy and lower vaccination coverage, however, studies found little to no negative impact on vaccine uptake in the US [30]. Additionally, moving the second dose of the measles, mumps, and rubella (MMR) vaccine to an earlier age was associated with improved vaccination coverage despite the schedule change in the UK [31]. Our findings may further contribute to the evidence that modifications to HPV vaccination schedules do not necessarily reduce vaccine uptake globally. Similar to our study finding, previous research has highlighted the difficulty of determining the extent to which changes in vaccination coverage can be attributed to a schedule change itself [32]. In our study, qualitative findings indicated that the single-dose switch was one of several factors shaping coverage trends, although we were unable to quantify its attributable contribution.

Our findings suggest that the COVID-19 pandemic was an important factor of global HPV vaccination coverage trends, particularly in non-HICs. Population-adjusted HPV1 coverage in non-HICs declined from 59.6% in 2019 to 45.7% in 2021 before recovering to 73.6% in 2024. This pattern aligns with global declines in childhood vaccination during the pandemic, with greater disruptions in lower-resource settings due to health system, program implementation, and economic vulnerabilities [33,34]. For HPV vaccination, high procurement and delivery costs and global supply constraints further limited vaccine access in many non-HICs [35]. Stakeholders in non-HICs described these supply-side challenges alongside vaccine hesitancy and reduced trust in governments. Pandemic-related disruptions to HPV vaccination programs were also reported in HICs, including those related to lockdowns, school closures and social distancing [36]. Consistent with these findings, stakeholders from Australia and the UK in our study, where school-based programs experienced coverage declines, identified reduced school attendance following the pandemic as an ongoing barrier. Nevertheless, population adjusted HPV vaccination coverage in HICs remained relatively stable, masking contextual variations across countries.

Recommendations to restore and improve HPV vaccination coverage in non-HICs have included greater global financial investment, adoption of single-dose schedules, expanded school-based delivery, and prioritization of single-age cohorts over MACs [35]. Previous studies found that stakeholders in non-HICs anticipated cost savings, reduced vaccine procurement needs, and improved vaccine stock management following a single-dose switch [16]. Our subgroup analyses showed significant increases in HPV1 coverage after the switch among non-HICs, countries in the African Region, Gavi-supported countries, and countries with school-based delivery, female-only programs, or single-age target cohorts. These overlapping subgroups share programmatic characteristics and include settings disproportionately affected by pandemic-related disruptions and supply constraints. Their coverage gains therefore may reflect the combined influence of schedule simplification, post-pandemic recovery, and efforts to address vaccine supply constraints.

Interviews across 15 countries further highlighted that coverage trends reflected multiple context-specific factors such as programmatic changes implemented alongside the switch. Several countries introduced MAC, gender-neutral vaccination, or routine HPV vaccination around the same time, consistent with previous reports suggesting that a single-dose schedule could serve as an enabler of these programmatic changes [17,37]. By reducing logistical demands and costs, a single-dose schedule may have facilitated the expansion of HPV vaccination program to broader target populations, particularly in non-HICs [15,38], potentially improving equity in vaccine access. However, a stakeholder’s account from Zambia and previous findings from Cameroon illustrate the difficulty of separating the effects of concurrent changes [20]. Thus, the observed coverage gains cannot be attributed solely to the schedule switch, although simplification may have enabled some of the accompanying program changes. Political and economic changes also influenced implementation and demand. Greater political stability has been associated with higher and more equitable childhood vaccination coverage than countries with lower political stability [39]. Consistent with this evidence, stakeholders in our study reported how governmental transitions affected resource allocation, staffing, vaccine procurement, supply chains, and program delivery. Stakeholders in Argentina also noted that an economic recession following a governmental transition shifted public attention toward more immediate needs and away from preventive health services, potentially reducing demand for HPV vaccination.

Several limitations should be noted. First, our quantitative analyses relied on annual country-level WUENIC estimates, which may be influenced by variations in reporting quality, completeness, and national data systems as reported in the qualitative interviews. We also could not account for countries that implemented the switch partway through a calendar year or only in selected regions. As a result, switch years were classified entirely as post-switch, potentially obscuring within-year and subnational variation in implementation. The findings should be interpreted as descriptive trends rather than estimates attributable to the switch itself. Second, despite repeated recruitment efforts, including up to four contact attempts for each identified stakeholder, the qualitative component included an average of 2.2 participants per country (range: 1-4). Given the small number of participants within individual countries, their perspectives may not fully reflect the range of experiences with HPV vaccination policy and implementation in each setting. Despite these limitations, to our knowledge, this is among the first studies to provide a global assessment of the switch to a single-dose HPV vaccination schedule across multiple countries and income settings. By integrating coverage analyses from 49 countries with stakeholder perspectives from 15 countries, we were able to contextualize quantitative findings and provide insights into factors underlying observed coverage changes.

## Conclusion

Our findings suggest that the switch to a single-dose schedule was associated improved coverage globally, with significant increase among non-HICs. Schedule simplification may have supported program recovery following disruptions caused by the COVID-19 pandemic and global HPV vaccine supply shortages, while enabling concurrent programmatic changes, including the introduction of MAC and gender-neutral vaccination strategies. Together, these changes may have expanded opportunities to reach previously underserved populations and reduce inequities in vaccine access. As adoption of single-dose schedule expands, countries should strengthen ways to reach missed populations and sustain coverage gains. Future evaluations should examine how schedule simplification interacts with delivery strategies, resource allocation, and broader program progress to determine vaccination outcomes. Continued monitoring of coverage, equity, and program sustainability is essential to guide implementation and assess progress toward cervical cancer elimination.

## Data Availability

HPV vaccination data used in this study are publicly available from the World Health Organization (WHO) websites. The interview data generated and analyzed during the current study are not publicly available because they contain information that could compromise participant confidentiality but are available from the corresponding author on reasonable request and subject to applicable data-sharing regulations.

## Acknowledgements

This work would not be possible without the national and sub-national immunization stakeholders who shared their time and insights. We extend our gratitude to them. We thank Dr. Hiroki Akaba and Dr. Sarah Snidal for their support in data analysis. We also acknowledge collaborators from the HAPPI Consortium (JSI Research & Training Institute, Inc., Clinton Health Access Initiative, Jhpiego, PATH), several of whom offered suggestions of key informants to interview or offered feedback in the conceptualization and post-analysis interpretation of this work.

## Supporting information captions

**S1 Table.** Key characteristics of countries represented by key informant interviews.

**S2 Table.** Selected domains and questions from the key informant interviews

**S3 Figure.** Population-adjusted HPVc vaccination coverage from 2010 to 2024, grouped by World Bank income category

**S4 Figure.** Box plots of mean HPV1 coverage by HPV vaccination program characteristics before and after the switch

**S5 Figure**. Bubble chart showing mean HPV1 coverage before and after the switch to a single-dose schedule among high-income countries (HICs) and non-HICs

**S6 Figure**. Bubble chart showing mean HPVc coverage before and after the switch to a single-dose schedule among high-income countries (HICs) and non-HICs

