## Supporting Information for "From Policy Change to Program Impact: HPV Vaccination Coverage Following the Global Shift to a Single-Dose Schedule"

### **Supplementary File**

**S1 Table.** Key characteristics of countries represented by key informant interviews.

| Country name | Country classification | HPV vaccine introduction year | Switch Year | Pre- post- coverage trend |
| --- | --- | --- | --- | --- |
| Argentina | UMIC | 2011 | 2024 | Decrease |
| Australia | HIC | 2007 | 2023 | Decrease |
| Brazil | UMIC | 2013 | 2024 | Stable |
| Cameroon | Gavi-supported LMIC | 2020 | 2023 | Increase |
| Canada | HIC | 2009 | 2024 | Stable |
| Colombia | UMIC | 2012 | 2023 | Increase |
| Ethiopia | Gavi-supported LIC | 2018 | 2024 | Stable |
| Lao PDR | Gavi-supported LMIC | 2020 | 2024 | Increase |
| Malaysia | UMIC | 2010 | 2024 | Increase |
| Mexico | UMIC | 2012 | 2022 | Increase |
| Peru | UMIC | 2015 | 2023 | Increase |
| Sierra Leone | Gavi-supported LIC | 2022 | 2023 | Decrease |
| South Africa | UMIC | 2014 | 2024 | Increase |
| United Kingdom | HIC | 2008 | 2023 | Decrease |
| Zambia | Gavi-supported LMIC | 2019 | 2023 | Stable |

Note: At the time of study, only two provinces in Canada (i.e., Quebec and British Columbia) adopted a switch to a single-dose schedule.

Abbreviations: Lao PDR: Lao People’s Democratic Republic, HIC: High-income country; UMIC: Upper-middle-income country; LMIC: Lower-middle-income country; LIC: Low-income country.

**S2 Table.** Selected domains and questions from the key informant interviews

| **Domains** | **Key questions** |
| --- | --- |
| Timing of the switch to a single-dose schedule and additional changes | Can you tell me about your position and work with the HPV vaccination program in your country?  When did the country switch to a single-dose schedule?  Were there any other changes made to the HPV vaccination program? |
| Coverage findings | What is your reaction to the change in coverage after the switch to a single-dose schedule?  What factors do you think influenced the observed coverage trend? |

**S3 Figure.** Population-adjusted HPVc vaccination coverage from 2010 to 2024, grouped by World Bank income category

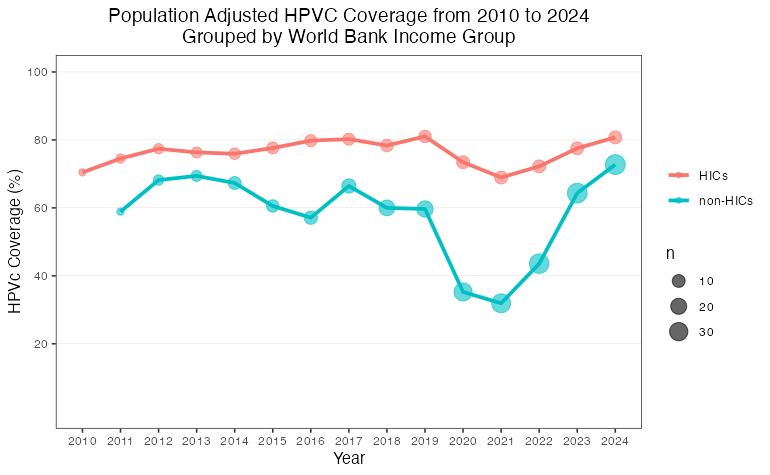

| **Income classification** | **2010** | **2011** | **2012** | **2013** | **2014** | **2015** | **2016** | **2017** | **2018** | **2019** | **2020** | **2021** | **2022** | **2023** | **2024** |
| --- | --- | --- | --- | --- | --- | --- | --- | --- | --- | --- | --- | --- | --- | --- | --- |
| HIC (%) | 70.5 | 74.5 | 77.4 | 76.3 | 75.9 | 77.6 | 79.8 | 80.2 | 78.4 | 81.0 | 73.4 | 68.9 | 72.2 | 77.5 | 80.8 |
| n | 3 | 4 | 5 | 6 | 7 | 8 | 8 | 8 | 9 | 9 | 10 | 10 | 10 | 10 | 10 |
| Non-HICs (%) |  | 58.9 | 68.2 | 69.4 | 67.3 | 60.6 | 57.1 | 66.4 | 60.0 | 59.7 | 35.3 | 31.9 | 43.6 | 64.3 | 72.7 |
| n | 0 | 3 | 5 | 6 | 10 | 9 | 11 | 14 | 19 | 21 | 29 | 33 | 36 | 37 | 38 |

Note: Implausible coverage estimates (i.e., 99%) were excluded from the analysis. Country income classifications were based on the 2025 World Bank income categories. Non-high-income countries (HICs) include low-income countries (LICs), lower-middle-income countries (LMICs), and upper-middle-income countries (UMICs).

**S4 Figure.** Box plots of mean HPV1 coverage by HPV vaccination program characteristics before and after the switch
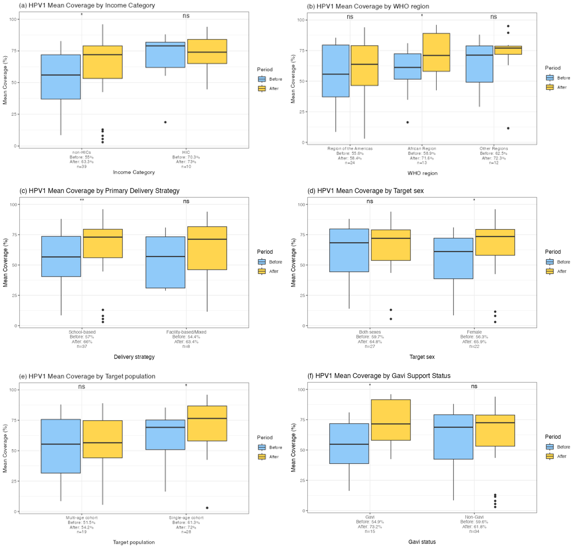

**S5 Figure**. Bubble chart showing mean HPV1 coverage before and after the switch to a single-dose schedule among high-income countries (HICs) and non-HICs

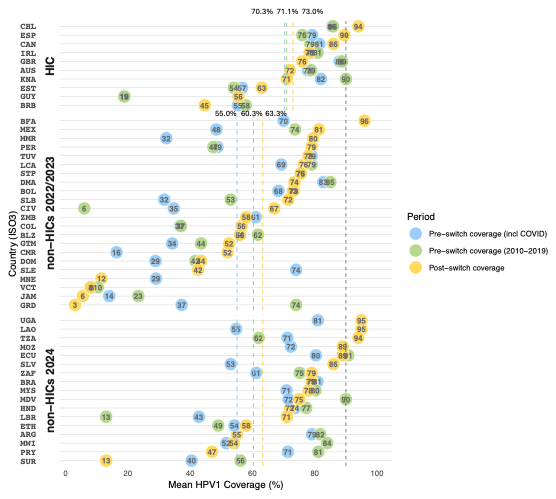

Note: Implausible coverage estimates (i.e., 99%) were excluded from the analysis. Countries were grouped into high-income countries (HICs) and non-HICs, as well as by the year of the switch (2022–2023 vs. 2024). Country names in the figure are presented using ISO Alpha-3 country codes. Both blue and green bubbles indicate coverage before the switch. Blue bubbles represent pre-switch coverage including the COVID-19 pandemic period, whereas green bubbles represent pre-switch coverage excluding the COVID-19 pandemic period. The dotted black line represents the WHO target of 90% HPV vaccination coverage. The colored dotted lines indicate the mean coverage in HICs and non-HICs.

**S6 Figure**. Bubble chart showing mean HPVc coverage before and after the switch to a single-dose schedule among high-income countries (HICs) and non-HICs

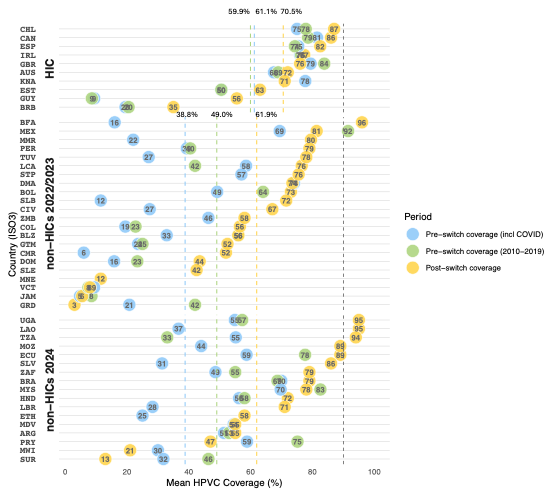

Note: Implausible coverage estimates (i.e., 99%) were excluded from the analysis. Countries were grouped into high-income countries (HICs) and non-HICs, as well as by the year of the switch (2022–2023 vs. 2024). Country names in the figure are presented using ISO Alpha-3 country codes. Both blue and green bubbles indicate coverage before the switch. Blue bubbles represent pre-switch coverage including the COVID-19 pandemic period, whereas green bubbles represent pre-switch coverage excluding the COVID-19 pandemic period. The dotted black line represents the WHO target of 90% HPV vaccination coverage. The colored dotted lines indicate the mean coverage in HICs and non-HICs. The mean values differ between HPV1 and HPVc because some countries reported different HPV1 and HPVc coverage in the switch year.
